# Methods used to select results to include in meta-analyses of nutrition research: a meta-research study

**DOI:** 10.1101/2021.08.05.21261680

**Authors:** Raju Kanukula, Joanne E McKenzie, Lisa Bero, Zhaoli Dai, Sally McDonald, Cynthia M Kroeger, Elizabeth Korevaar, Matthew J Page

**Affiliations:** School of Public Health and Preventative Medicine, Monash University, 553 St Kilda Road, Melbourne, Victoria 3004, Australia; Center for Bioethics and Humanities, University of Colorado Anschutz Medical Campus, 13080 E. 19th Ave, Aurora, CO 80045, United States; Australian Institute of Health Innovation, Macquarie University, 75 Talavera Rd, North Ryde, NSW 2113, Australia; Charles Perkins Centre, School of Pharmacy, Faculty of Medicine and Health, The University of Sydney, D17, The Hub, 6^th^ Floor, Camperdown, NSW, 2006, Australia; Charles Perkins Centre, Central Clinical School, Faculty of Medicine and Health, The University of Sydney, D17, The Hub, 6^th^ Floor, Camperdown, NSW, 2006, Australia

**Keywords:** Meta-analysis, Systematic review, Nutrition, Multiplicity, Eligibility criteria, Decision rule

## Abstract

**Objectives:** To investigate the extent of multiplicity of results in study reports of nutrition research, and the methods specified in systematic reviews to select results for inclusion in meta-analyses.

**Methods:** MEDLINE and Epistemonikos were searched (January 2018 – June 2019) to identify systematic reviews with meta-analysis of the association between food/diet and health-related outcomes. A random sample of these reviews was selected, and for the first presented (‘index’) meta-analysis, rules used to select effect estimates to include in this meta-analysis were extracted from the reviews and their protocols. All effect estimates from the primary studies that were eligible for inclusion in the index meta-analyses were extracted.

**Results:** Forty-two systematic reviews were included, 14 of which had a protocol. In 29% of review protocols and 69% of reviews, at least one decision rule to select effect estimates when multiple were available was specified. In 69% (204/325) of studies included in the index meta-analyses, there was at least one type of multiplicity of results.

**Conclusions:** Authors of systematic reviews of nutrition research should anticipate encountering multiplicity of results in the included primary studies. Specification of methods to handle multiplicity when designing reviews is therefore recommended.

What is new?

Key Findings

- Authors of systematic reviews of nutrition research should anticipate encountering multiplicity of results in the included primary studies.
- Decision rules to select results for inclusion in meta-analyses of nutrition research were infrequently pre-specified.

What this adds to what was known?

- Previous studies have found that multiplicity of results of continuous outcomes in studies included in systematic reviews was common, and methods used to select results to include in meta-analyses were infrequently pre-specified in systematic review protocols. However, none of the studies examined meta-analyses in nutiriton research, inclusion of randomized or non-randomized studies, or where the outcome was non-continuous (e.g. binary, count or time-to-event); circumstances for which different forms of multiplicity might arise. Our study addressed this gap.

What is the implication and what should change now?

- Pre-specification of decision rules to handle multiplicity when designing reviews is recommended. In the systematic review, we recommend reporting any modifications to the specified rules, or any additions that were introduced to cover multiplicity scenarios that had not been anticipated when designing the review.

## 1. Background

The Global Burden of Disease study 2019 reported that diet has a significant impact on health outcomes. Diet quality was found to be the fifth leading risk factor for disability adjusted life years [1]. Large and long-term prospective observational studies and short-term clinical trials have found associations between a particular dietary factor and non-communicable diseases [2-4]. Systematic reviews (SRs) based on such studies are being used to inform recommendations in dietary guidelines [5-7]. However, flaws in the design, conduct and reporting of SRs may yield misleading results, and in turn, misinform guideline recommendations [8].

One challenge SR authors often face is a multiplicity of results in the primary studies [9]. For example, a study report may present multiple effect estimates for the association between red meat consumption and gout, where these estimates may arise from the fitting of multiple statistical models with different outcome definitions of gout, different exposure levels, or where adjustment is made for different sets of confounders. While inclusion of multiple effect estimates from a particular study in a meta-analysis is possible (using methods that adjust for statistical dependency) [10], more commonly only one of the available effect estimates is selected for inclusion. There are various methods that can be used to select a single effect estimate [9, 10]. However, when this selection is based on the statistical significance, magnitude or direction of effect, this may introduce bias into the meta-analysis effect estimate [11]. We refer to this selection process as ‘selective inclusion of results’.

To help mitigate the potential for selective inclusion of results, it has been recommended that methods for dealing with multiplicity should be pre-specificied [11]. This includes pre-specification of “eligibility criteria” for each meta-analysis, indicating which results are eligible for inclusion in the meta-analysis (e.g. intervention groups, measurement instruments, time points), and “decision rules”, which specify the methods that will be used to select a single result from a study when multiple are eligible for inclusion in the same meta-analysis (see Box 1 for examples of eligibility criteria and decision rules).

### Box 1.

**Examples of eligibility criteria and decision rules to select results**

*Example of eligibility criteria to select results:* Systematic reviewers state that only study effect estimates that were adjusted for sex and age would be included in a meta-analysis of the association between fruit consumption and coronary heart disease.

*Example of a decision rule to select results when multiple are available:* Systematic reviewers state that if multiple effect estimates quantifying the association between different levels of fruit consumption and coronary heat disease were available, as would arise if intake was categorised into quartiles in a study report, only the contrast between the highest (e.g. quartile 4) and lowest (e.g. quartile 1) intake would be included in the meta-analysis.

Previous research has examined the extent of multiplicity of results, and the methods used to select results for inclusion in the meta-analysis [11-13]. These studies have focused on a range of conditions and examined multiplicity in meta-analyses of randomised trials with continuous outcomes. All studies found that multiplicity of results was common, and Page et al.[9] and Tendal et al. [12] found that specification of methods to select results for inclusion were rarely reported. However, none of the studies examined meta-analyses in nutiriton research, inclusion of randomized or non-randomized studies, or where the outcome was non-continuous (e.g. binary, count or time-to-event); circumstances for which different forms of multiplicity might arise. Therefore, we aimed to address this gap and investigate the i) extent of multiplicity of results in sudy reports of nutrition research, and ii) the methods specified in systematic reviews to select results for inclusion in meta-analyses of all outcome types, including randomized or non-randomized study designs.

## 2. Methods

The study protocol has been published [14]. Here, we provide an overview of the methods, with modifications to the protocol reported in Supplementary Table 1.

### 2.1. Eligibility criteria, search and selection of SRs

SRs that satisfied the definition of an SR, as outlined in the 2019 edition of the Cochrane Handbook for Systematic Reviews of Interventions [15], and that had explicitly stated methods of study identification (e.g. a search strategy) and of study selection (e.g. eligibility criteria and selection process), and included a meta-analysis of study results, were eligible for inclusion in this study. We included such SRs with meta-analysis that:

- included studies that enrolled, regardless of their age and background, (a) people who were generally healthy, (b) a mixture of generally healthy people and people with diet-related risk factors (e.g. overweight, high blood pressure) or a particular health condition (e.g. type II diabetes or cardiovascular disease), or (c) people with non-specified health status;
- included randomized trials or non-randomized studies that assessed the effects of at least one type of food (e.g. eggs, fish) or at least one food-defined dietary pattern (e.g. Mediterranean diet) on any continuous (e.g. systolic blood pressure) or non-continuous (e.g. gout incidence) health-related outcome;
- were published between 1 January 2018 and 30 June 2019;
- were written in English;
- provided citations for all included studies in the SR, and;
- presented the summary statistics or effect estimate and its precision (e.g. standard error or 95% confidence interval) for each included study, and the meta-analytic summary effect estimate and its precision in the text or forest plot, for at least one meta-analysis of a continuous or non-continuous outcome.

We excluded

- SRs that did not include any meta-analysis of a non-continuous or continuous outcome;
- meta-analyses or pooled analyses of studies conducted outside the context of a SR;
- SRs that only focused on nutrient-specific associations with outcomes (e.g. examining the effects of single nutrients such as folic acid, salt);
- SRs that included studies enrolling only participants with a health condition, who were obese or who were frail or elderly people at risk of malnutrition, and;
- SRs that were co-authored by any of our research team members.

We searched for eligible SRs indexed in the PubMed and Epistemonikos [16] databases from 1 January 2018 to 30 June 2019 (search strategies reported in supplement 2). The search results were exported into Microsoft Excel, all duplicate records were removed, and the remaining records were randomly sorted. In the piloting phase, four investigators (MJP, CMK, ZD and SM) independently assessed 50 abstracts against the inclusion criteria (rating each as ‘Eligible’, ‘Ineligible’, or ‘Unsure’), discussed any discrepancies, and made any necessary changes to the screening form. Following piloting, two investigators (MJP and one of CMK, ZD or SM) independently screened titles and abstracts of 450 records. Two investigators (MJP and one of CMK, ZD or SM) then independently assessed the full text of records that were rated as ‘Eligible’ or ‘Unsure’ against the eligibility criteria. This screening process was repeated (in batches of 500 records) until we reached the target sample of 50 SRs, including 25 meta-analyses of continuous and 25 meta-analyses of non-continuous outcomes. If the total number of eligible SRs exceeded this target at the end of a batch, we planned to randomly sample 25 SRs of each type. Any discrepancies in screening decisions at each stage were resolved via discussion between investigators, or by consultation with another investigator (JM) where necessary. For each included SR, we retrieved the published protocol for the SR or registration record (e.g. PROSPERO record), if available, as cited or reported in the SR.

From each SR meeting the inclusion criteria, one investigator (MJP) selected one meta-analysis for inclusion. The selected meta-analysis was the first meta-analytic result mentioned in the review (with no restrictions on its placement in the manuscipt); we refer to the selected meta-analysis as the “index meta-analysis”. We initially selected an index meta-analysis regardless of the outcome domain (e.g. weight, bladder cancer), effect measure (e.g. odds ratio, standardised mean difference), meta-analytic model and number and type of included studies (i.e. randomized or non-randomized study). However, following selection of 50 index meta-analyses, we decided to restrict our inclusion to only those meta-analyses including fewer than 20 studies, for reasons of feasibility. For each included index meta-analysis, we retrieved the reports of all included studies (see the protocol for further details [14]).

### 2.2 Data collection and management

A data collection form was developed in REDCap (see Supplementary Table S2) [17]. Seven investigators (RK, ZD, SM, CMK, EK, LB and MJP) piloted the form on two randomly selected meta-analyses and their included studies. Discrepancies were discussed, and we modified the form accordingly. Following piloting, two investigators (RK and one of ZD, SM, CMK, LK, LB and MJP) independently collected data from a random sample of half of the index meta-analyses and their included studies, while one investigator (RK) collected data on the remaining index meta-analyses and their included studies. Any discrepancies were resolved through discussions between two investigators or adjudication by a third investigator (JM) if necessary.

An overview of the data items, and the sources these were obtained from (i.e. systematic review protocol, systematic review or study report), is presented in Table 1; further details are available in the protocol [14]. In the case of data extracted from the reports of studies included in the index meta-analysis, we extracted all outcome data that were eligible for inclusion in the index meta-analysis. This was determined by the eligibility criteria and decision rules stated in the SR protocol if available, and if not available, the SR, and in combination with the comparison and outcome of the meta-analysis. For example, if the systematic reviewers pre-defined in the SR protocol that the eligible intervention and comparator for the meta-analysis of weight gain was “highest versus lowest intake of dairy products”, and pre-defined a decision rule stipulating that they would consider only data at 12 weeks follow-up when data were available at multiple time points, we only extracted data for that comparison and time point, regardless of whether study reports had data for other time points and other comparisons for the same outcome. In the absence of an SR protocol, we assumed that no eligibility criteria and decision rules were pre-specified (‘worst-case scenario’ assumption) and extracted all study outcome data based on how the outcome was specified in the SR. For example, if the systematic reviewers did not state in a protocol which results should be selected when multiple were available, yet defined the meta-analysis as “effect of dairy intake on weight at 6 months”, we extracted all data on weight at 6 months, regardless of the level of intake of dairy, whether results were unadjusted or adjusted, or what analysis sample was used.

**Table 1:** Data sources and data items (see protocol for further details) [14].

| Source | Data items |
| --- | --- |
| Systematic review protocol | Year of publication/registration; eligibility criteria and decision rules to select results to include in the index meta-analysis |
| Systematic review | *General characteristics of the systematic review*<br>Journal name; year of publication; corresponding author's country and affiliation; conflicts of interest of review authors; source of funding for the review;<br>*General characteristics of the index meta-analysis*<br>Number of studies and participants; type of population investigated; type of interventions/exposures investigated; type of studies included in the meta-analysis; outcome domain (such as weight, cardiovascular function); outcome primacy label (primary or secondary or unlabelled); meta-analysis effect measure; meta-analysis model; eligibility criteria and decision rules to select results to include in the index meta-analysis; summary statistics, effect estimates and measures of precision (e.g. confidence interval) for each included study; and the meta-analytic effect estimate and measure of precision. |
| Study reports | *Outcome data that could potentially be included in the index meta-analysis*<br>Outcome definition and measurement instrument; intervention/exposure description; comparator description; time point; analysis sample (e.g. intention-to-treat, per-protocol); summary statistics (e.g. number of events and sample sizes of both intervention/exposure and comparator); effect measure (e.g. risk ratio, mean difference); effect estimates and measures of precision (e.g. 95% confidence interval) and location of data in the report; whether results were unadjusted or adjusted for covariates, covariates that were adjusted for (if applicable). |

### 2.3 Data analysis

We used descriptive statistics to summarise the characteristics of SR protocols, SRs, index meta-analyses and included studies. For categorical variables, we present frequencies and percentages. For continuous variables, we report medians (with interquartile ranges [IQR]). We computed the frequencies and percentages for different types of eligibility criteria and decision rules used to select results, differences in eligibility criteria or decision rules between the SR protocol and the SR, and studies with different types of multiplicity of results. We also calculated risk differences (with 95% confidence intervals [CIs]) to examine whether the percentages of different types of eligibility criteria and decision rules and studies with different types of multiplicity of results, differed between meta-analyses of continuous outcomes and of non-continuous outcomes. Risk differences were calculated using the method of Mee with the Miettinen and Nurminen modification. Analyses were undertaken using the statistical packages Stata (College Station, Tx), except for the calculaton of risk differences, which was undertaken using the library PropCIs [18] in R (Vienna, Austria) [19].

## 3. Results

### 3.1 Results of search and screening

Our search yielded a total of 7,167 references from the PubMed and Epistemonikos databases (Figure 1). After removing duplicates (n=908), we screened the titles and abstracts of 3,013 randomly sorted references, of which 2,777 were excluded, leaving 236 for full-text screening. Of these, 99 SRs met the inclusion criteria, including 25 SRs with a meta-analysis of a continuous outcome and 74 with meta-analysis of a non-continuous outcome. Initially, all SRs with a continuous outcomes were included, and 25 of the SRs with a non-continuous outcome were randomly selected. From these 50 SRs, eight were excluded (6 continuous, 2 non-continuous), because the index meta-analysis had 20 or more studies, leaving 42 included SRs [20-61].

**Figure 1.**
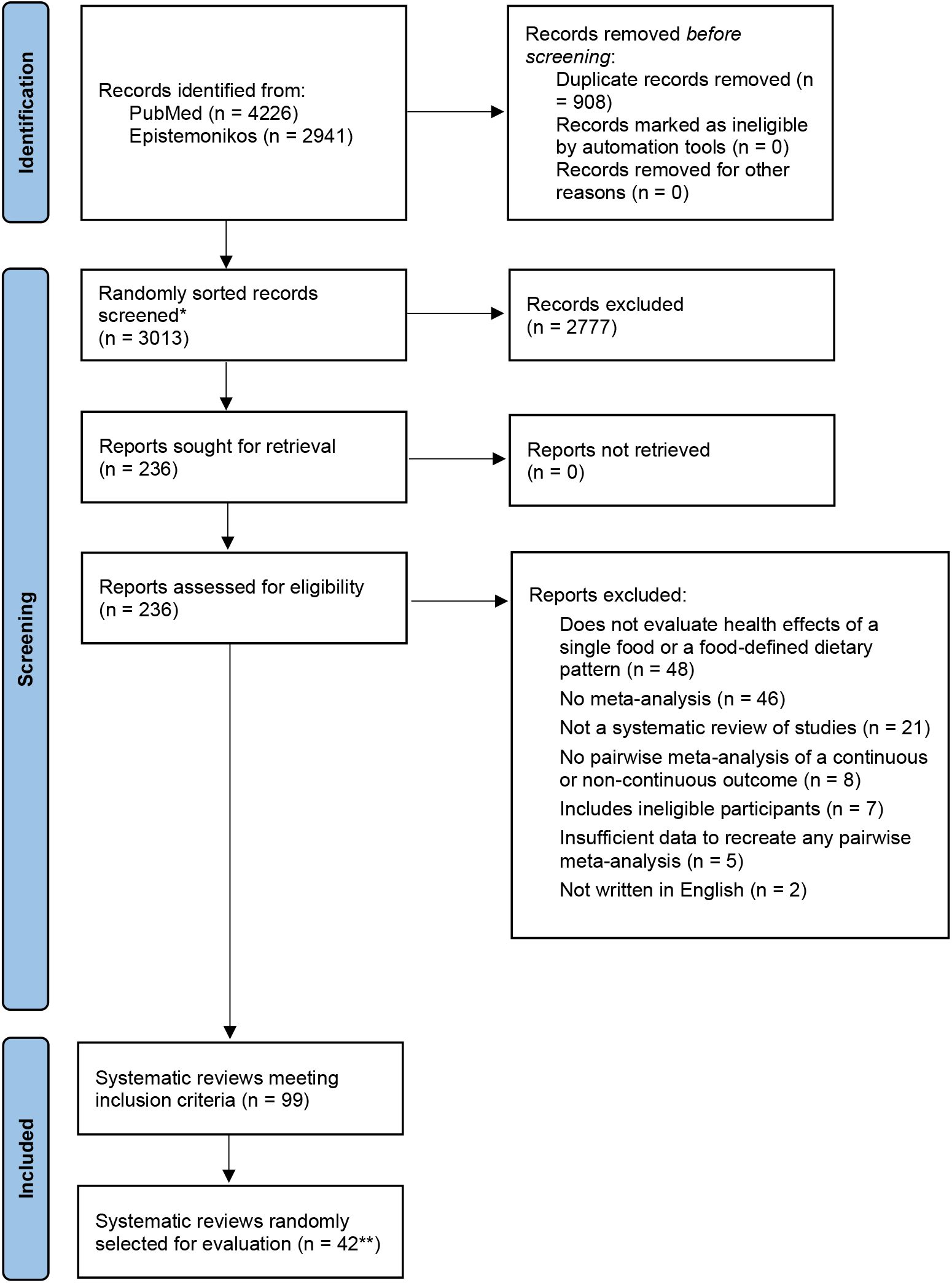
PRISMA 2020 flow diagram of identification, screening and inclusion of systematic reviews. *Of the 6259 unique titles and abstracts, we only needed to screen 3,013 randomly sorted titles and abstracts to reach our target sample size. **We initially drew a random sample of 50 systematic reviews, but post-hoc excluded eight systematic reviews with 20 or more included studies in the index meta-analysis to reduce workload.

### 3.2 Characteristics of the included systematic reviews and their index meta-analyses

Of the 42 SRs, 14 had accessible protocols (2 were published protocols and 12 were PROSPERO records) (Table 2). In most index meta-analyses the population was unclear (45%, 19/42), 33% (14/42) included only healthy participants and the remainder (22%, 9/42) included a mix of healthy people and people with a health condition. The majority of index meta-analyses included only non-randomized studiess (62%, 26/42), 31% (13/42) included only randomized trials and the remaining 7% included studies of both designs (Table 3). The primacy of the outcome was not identified in most reviews (81%, 34/42). In nearly all meta-analyses, a random-effects model was fitted (90%, 38/42). The 42 index meta-analyses included a total of 325 studies, with a median of seven studies (IQR 5-11; range 2-17) per meta-analysis.

**Table 2:** Characteristics of the systematic reviews (N=42)

| **Characteristic** | **n (%)** |
| --- | --- |
| Focus of journal |  |
| Restricted to nutrition research | 28 (67) |
| Not restricted to nutrition research | 14 (33) |
| Country of the corresponding author(s) |  |
| China | 9 (21) |
| Iran | 7 (17) |
| United States of America | 6 (14) |
| Others (Australia, Austria, Brazil, Canada, Israel, Japan, Malaysia, Spain, Sweden, Thailand, United Kingdom) | 20 (48) |
| Affiliation of the corresponding author(s) |  |
| Food industry | 2 (5) |
| Non-industry | 37 (88) |
| Mixed | 2 (5) |
| Unclear | 1 (2) |
| Source of funding |  |
| Non-profit | 23 (55) |
| For-profit | 3 (7) |
| Mixed | 0 |
| No funding | 8 (19) |
| Not reported | 8 (19) |
| Conflict of interest |  |
| Conflict of interest reported by at least one review author | 7 (17) |
| All review authors stated they had no conflicts of interest | 29 (69) |
| Missing/not reported | 6 (14) |
| Protocol availability |  |
| Both a protocol and registration record are available | 0 |
| Only a protocol is available | 2 (5) |
| Only a registration record is available | 12 (29) |
| Neither are available | 28 (67) |
| Protocol published year* | 2012 & 2017 |
| Protocol registered year (median, [IQR]) | 2018 (2017-2018) |

\*Only two protocols published

**Table 3:** Characteristics of index meta-analyses (N=42)

| **Characteristics** | **n (%)** |
| --- | --- |
| Type of participants in included studies |  |
| Healthy only | 14 (33) |
| Mix of healthy people and people with a health condition | 9 (21) |
| Unclear | 19 (45) |
| Type of included studies |  |
| Only randomised trials | 13 (31) |
| Only non-randomised trials | 26 (62) |
| Both | 3 (7) |
| Total number of studies included (median [IQR]) | 7 (5-11) |
| Total number of participants (median [IQR]) | 2972 (857-44418)* |
| Outcome labelling |  |
| Primary | 6 (14) |
| Secondary | 2 (5) |
| Unlabelled | 34 (81) |
| Outcome type |  |
| Continuous | 19 (45) |
| Non-continuous (e.g. binary, count, time-to-event) | 23 (55) |
| Meta-analytic effect measure |  |
| Risk ratio | 15 (36) |
| Odds ratio | 6 (14) |
| Hazard ratio | 2 (5) |
| Mean difference | 18 (43) |
| Standardised mean difference | 1 (2) |
| Index meta-analysis model used |  |
| Fixed-effect | 2 (5) |
| Random-effects | 38 (90) |
| Unclear | 2 (5) |
| Type of intervention/exposure |  |
| Dairy | 5 (12) |
| Fruit | 2 (5) |
| Pescatarian | 2 (5) |
| Vegan | 1 (2) |
| Vegetarian | 12 (28) |
| Mediterranean diet | 5 (12) |
| Non-veg | 1 (2) |
| Chocolate | 2 (5) |
| Mixed** | 12 (28) |

\* Only 17 of the 42 SRs reported the total number of participants included in the index meta-analysis
\*\* Includes combinations of fruits, meat, egg, milk, fish and vegetables etc.

### 3.3 Eligibility criteria and decision rules reported in SR protocols

Of the SR protocols (n=14), all included at least one eligibility criterion, and four (29%) reported at least one decision rule to select results (Table 4). Almost all protocols specified eligibility criteria based on interventions/exposures (93%, 13/14) (e.g.specifying which foods or dietary patterns were eligible), but few other types of eligibility criteria were specified (e.g. time points [29%, 4/14], information sources [13%, 3/14]). The most commonly pre-specified decision rule was based on interventions/exposures (reported in 3 of the 4 SR protocols with at least one decision rule to select results). See Supplementary Table S3 for the content of the decision rules.

**Table 4.** Number of systematic review protocols or registration entries reporting eligibility criteria and decision rules to select results (N=14)

| **Criteria** | **Total**<br>**n (%)**<br>**[n=14]** | **SRs of**<br>**continuous**<br>**outcomes**<br>**n (%)**<br>**[n=9]** | **SRs of non-**<br>**continuous**<br>**outcomes**<br>**n (%)**<br>**[n=5]** | **Risk difference\***<br>**95% confidence**<br>**interval\*\*** |
| --- | --- | --- | --- | --- |
| Total |  |  |  |  |
| At least one eligibility criterion | 14 (100) | 9 (100) | 5 (100) | 0 (-31, 45) |
| At least one decision rule | 4 (29) | 2 (22) | 2 (40) | -18 (-62, 30) |
| Measurement instruments |  |  |  |  |
| Eligibility criteria | 1 (7) | 1 (11) | 0 | 11 (-36, 45) |
| Decision rule | 0 | 0 | 0 | 0 (-45, 31) |
| Definitions/diagnostic criteria |  |  |  |  |
| Eligibility criteria | 0 | 0 | 0 | 0 (-45, 31) |
| Decision rule | 0 | 0 | 0 | 0 (-45, 31) |
| Cut-points on a measurement instrument |  |  |  |  |
| Eligibility criteria | 0 | 0 | 0 | 0 (-45, 31) |
| Decision rule | 0 | 0 | 0 | 0 (-45, 31) |
| Time points |  |  |  |  |
| Eligibility criteria | 4 (29) | 2 (22) | 2 (40) | -18 (-63, 30) |
| Decision rule | 0 | 0 | 0 | 0 (-45, 31) |
| Interventions/exposures |  |  |  |  |
| Eligibility criteria | 13 (93) | 9 (100) | 4 (80) | 20 (-16, 63) |
| Decision rule | 3 (21) | 2 (22) | 1 (20) | 2 (-47, 43) |
| Information sources |  |  |  |  |
| Eligibility criteria | 3 (21) | 2 (22) | 1 (20) | 2 (-47, 43) |
| Decision rule | 1 (7) | 0 | 1 (20) | -20 (-63, 16) |
| Analyses |  |  |  |  |

| Criteria | Total<br>n (%)<br>[n=14] | SRs of<br>continuous<br>outcomes<br>n (%)<br>[n=9] | SRs of non-continuous<br>outcomes<br>n (%)<br>[n=5] | Risk difference*<br>95% confidence<br>interval** |
| --- | --- | --- | --- | --- |
| Eligibility criteria for any type of analysis | 2 (14) | 2 (22) | 0 | 22 (-27, 56) |
| Decision rule for any type of analysis | 2 (14) | 0 | 2 (40) | -40 (-78, 0) |
| Rule for final vs. change from baseline values | 0 | 0 | NA | NA |
| Rule for analyses undertaken on multiple samples (e.g., ITT vs. per-protocol) | 0 | 0 | 0 | 0 (-45, 31) |
| Rule for unadjusted vs. covariate-adjusted analyses | 1 (7) | 0 | 1 (20) | -20 (-63, 16) |
| Rule for period vs. paired analyses in crossover randomized trials | 0 | 0 | 0 | 0 (-45, 31) |
| Rule to handle results arising from overlapping samples of participants | 1 (7) | 0 | 1 (20) | -20 (-63, 16) |
| Other decision rule (Multiplicity) | 0 | 0 | 0 | 0 (-45, 31) |

\*Risk difference calculated as the difference in percentage of SRs reporting the specified eligibility criteria/decision rule between SRs of continuous outcomes minus SRs of non-continuous outcomes. ITT: Intention to treat; NA: Not applicable;
\*\*Confidence limits for the difference in percentages calculated using the method of Mee with the Miettinen and Nurminen modification

### 3.4 Eligibility criteria and decision rules reported in SRs

Of the SRs (n = 42), all included at least one eligibility criterion and 69% reported at least one decision rule to select results (Table 5). Similar to the SR protocols, the most commonly reported eligibility criteria (95%, 40/42) and decision rule (40%, 17/42) in the SRs was based on interventions/exposures. Eligibility criteria and decision rules for the type of analysis were more freqently specified in SRs as compared with their protocols. The most commonly reported decision rules for analyses were rules to select from multiple unadjusted and covariate-adjusted analyses (24%) (Table 5). There were some discrepancies observed between SR protocols and their published SRs, with the most common type being the addition of a new decision rule to deal with multiple unadjusted and covariate-adjusted analyses in the included studies (Supplementary Table S4).

**Table 5.** Number of systematic reviews reporting eligibility criteria and decision rules to select results.

| **Criteria** | **Total SRs**<br>**n (%)**<br>**[n = 42]** | **SRs of**<br>**continuous**<br>**outcomes**<br>**n (%)**<br>**[n = 19]** | **SRs of non-**<br>**continuous**<br>**outcomes**<br>**n (%)**<br>**[n = 23]** | **Risk difference\***<br>**95% Confidence**<br>**interval \*\*** |
| --- | --- | --- | --- | --- |
| Total |  |  |  |  |
| At least one eligibility criterion | 42 (100) | 19 (100) | 23 (100) | 0 (-17, 14) |
| At least one decision rule | 29 (69) | 14 (74) | 15 (65) | 8 (-20, 35) |
| Measurement instruments |  |  |  |  |
| Eligibility criteria | 1 (2) | 1 (5) | 0 | 5 (-9, 24) |
| Decision rule | 2 (5) | 1 (5) | 1 (4) | 1 (-17, 21) |
| Definitions/diagnostic criteria |  |  |  |  |
| Eligibility criteria | 2 (5) | 0 | 2 (9) | -9 (-27, 9) |
| Decision rule | 1 (2) | 0 | 1 (4) | -4 (-21, 13) |
| Cut-points on a measurement instrument |  |  |  |  |
| Eligibility criteria | 0 | 0 | 0 | 0 (-15, 17) |
| Decision rule | 0 | 0 | 0 | 0 (-15, 17) |
| Time points |  |  |  |  |
| Eligibility criteria | 4 (10) | 2 (11) | 2 (9) | 2 (-18, 24) |
| Decision rule | 4 (10) | 3 (16) | 1 (4) | 11 (-8, 34) |
| Interventions/exposures |  |  |  |  |
| Eligibility criteria | 40 (95) | 19 (100) | 21 (91) | 9 (-9, 27) |
| Decision rule | 17 (40) | 6 (32) | 11 (48) | -16 (-43, 14) |
| Information sources |  |  |  |  |
| Eligibility criteria | 3 (7) | 1 (5) | 2 (9) | -3 (-23, 17) |
| Decision rule | 4 (10) | 3 (16) | 1 (4) | 11 (-8, 34) |
| Analyses |  |  |  |  |
| Eligibility criteria for any type of analysis | 11 (26) | 7 (37) | 4 (17) | 20 (-7, 45) |

| Criteria | Total SRs<br>n (%)<br>[n = 42] | SRs of<br>continuous<br>outcomes<br>n (%)<br>[n = 19] | SRs of non-continuous<br>outcomes<br>n (%)<br>[n = 23] | Risk difference*<br>95% Confidence<br>interval ** |
| --- | --- | --- | --- | --- |
| Decision rule for any type of analysis | 16 (38) | 8 (42) | 8 (35) | 7 (-21, 35) |
| Rule for final vs. change from baseline values | 3 (7) | 3 (16) | 0 | NA |
| Rule for analyses undertaken on multiple samples (e.g.,<br>ITT vs. per-protocol) | 0 | 0 | 0 | 0 (-15, 17) |
| Rule for unadjusted vs. covariate-adjusted analyses | 10 (24) | 2 (11) | 8 (35) | -24 (-47, 2) |
| Rule for period vs. paired analyses in crossover<br>randomized trials | 2 (5) | 2 (11) | 0 | 11 (-5, 31) |
| Rule to handle results arising from overlapping samples of<br>participants | 1 (2) | 0 | 1 (4) | -4 (-21, 13) |
| Other decision rule | 5 (12) | 1 (5) | 4 (17) | -12 (-33, 10) |

\*Risk difference calculated as the difference in percentage of SRs reporting the specified eligibility criteria/decision rule between SRs of continuous outcomes minus SRs of non-continuous outcomes. ITT: Intention to treat; NA: Not applicable;
\*\*Confidence limits for the difference in percentages calculated using the method of Mee with the Miettinen and Nurminen modification

The percentage of reviews specifying different types of eligibility criteria and decision rules generally did not differ by outcome type. However, a larger percentage of SRs with an index meta-analysis of a continuous outcome specified eligibility criteria for any type of analysis compared with SRs with a non-continuous outcome (37% vs 17%; risk difference [RD] 20%, 95% CI -7% to 45%). Conversley, a smaller percentage of SRs with an index meta-analysis of a continuous outcome specified a rule for selecting an adjusted/unadjusted result compared with SRs of a non-continuous outcome (11% vs 35%; RD -24%, 95% CI -47% to 2%).

### 3.5 Multiplicity of results in included studies

Of the 325 studies, 296 studies were included in reviews (n=38) without any prespecified decision rules to select results to include in meta-analyses (Table 6). These reviews, and the studies within, are therefore used to estimate the extent of multiplicity of results that can be expected when no decision rules to select results are predefined. The median (IQR) number of available effect estimates per study was 2 (1 to 3), and the largest number of effect estimates in a study was 41 [62]. The most common types of multiplicity arose from multiple unadjusted and one or more covariate-adjusted analyses (which occurred in 40% of the included studies), followed by multiple intervention/control groups (24%). The least common types of multiplicity arose from multiple instruments (0%). The studies with continuous outcomes had less multiplicity than the studies with non-continuous outcomes (54% vs 80%; RD -26%, 95% CI -40%, -12%).

**Table 6:**
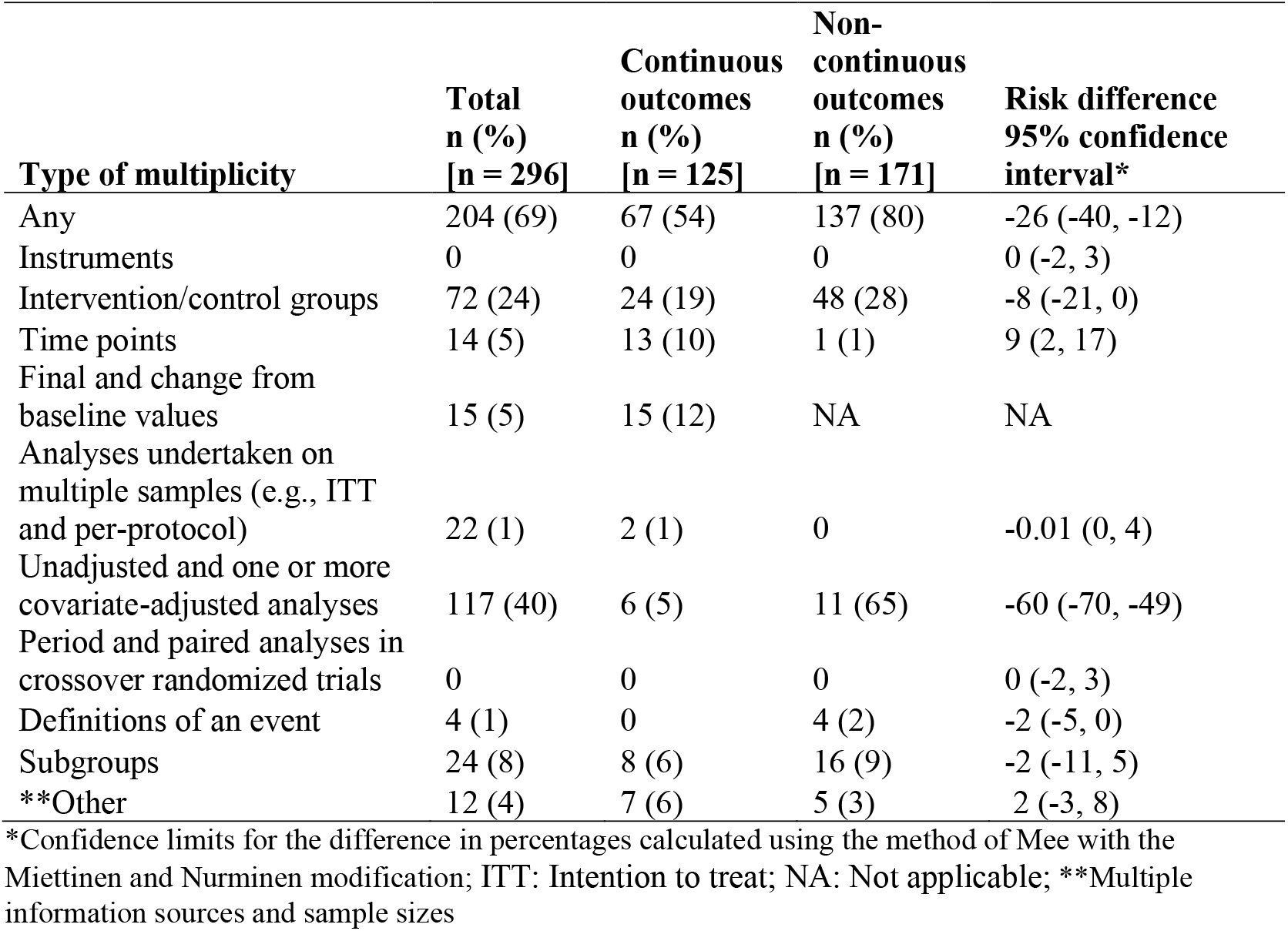
**Number of studies with different types of multiplicity of results in systematic reviews without prespecified methods to select results (N=38)**

## 4. Discussion

Our findings show that decision rules to select results were infrequently pre-specified in the protocols of a randomly selected sample of SRs with meta-analyses of nutrition research. Multiplicity of results in the primary studies included in the index meta-analyses was very common, with 69% having at least one type of multiplicity. The frequency and types of multiplicity in the included studies varied, arising from multiple intervention groups, time points, analyses and subgroups.

### Comparison with previous research

The findings of our study are in line with previously published studies, which have observed incomplete pre-specification of SR methods such as eligibility criteria, methods for collecting, handling and analysing data, and pre-specification of eligibility criteria and decision rules in PROSPERO records [63], Cochrane protocols [9, 64] and published SRs [9, 65]. Two previous studies, on which the methodology of the present study is based, examined methods used to select results for inclusion in meta-analyses [12] [9]. Page et al. [9] examined 44 SRs,, nearly half of which were Cochrane reviews, and they included only randomised trials. In Page et al. [9] and our study, all protocols reported at least one eligibility criteria, but at least one decision rule was pre-specified more in the SR protocols included in Page et al. [9] than those in our study (81% vs 29%). The frequency of discrepancies between the SR protocol and SR in the eligibility criteria or decision rules to select results was higher in our study. Tendal et al. [12] examined eighteen Cochrane reviews, all of which had protocols. Eight of the protocols mentioned eligible time points or periods, but only one provided decision rules to handle multiplicity of time points. Interestingly, all of the 18 protocols reported eligibility criteria for the control group, but none reported decision rules to handle multiple control groups in included studies.

Three studies (Tendal et al.[12], Mayo-Wilson et al. [13] and Page et al.[9]) assessed the multiplicity of results among the included studies of SRs. Compared to our study, Page et al. [9] and Tendal et al. [12] had fewer studies with multiple estimates that were available for inclusion in a particular meta-analysis. Multiplicity arising from multiple intervention/control groups was slightly less frequent in our study compared to Tendal et al. [12] (24% vs 29%) but was more frequent in ours when compared with Page et al. [9] (24% vs 17%). Our study findings showed less multiplicity in terms of time points (5%) and measurement instruments for outcomes (0%) compared to previous studies [9, 12]. However, we observed greater multiplicity (40%) due to unadjusted and one or more covariate-adjusted analyses. This difference was likely driven by inclusion of non-randomized studies in the present study, which often require adjustment for covariates to reduce risk of bias due to confounding [66]. Similarly, Mayo-Wilson et al. [13] assessed multiplicity in the clinical trials of publicly accessible reports and non-public reports related to gabapentin for neuropathic pain (n=21) and quetiapine for bipolar depression (n=7). In 15/21 (71%) gabapentin trials and 7/7 (100%) quetiapine trials, there was multiplicity of results.

### Strengths and limitations

The major strength of our study is that we have pre-specified methods to identify, select and collect data from eligible SRs and studies, and provided the modifications/deviations to our study protocol (supplementary Table S1). We also used extensive search strategies to identify SRs in nutrition research. In addition, all the study authors, who had different levels of expertise, undertook training and pilot-testing of data collection forms. Moreover, given we randomly selected the SRs, our findings are generalisable to SRs meeting this study’s eligibility criteria.

A limitation of our study is that we only retrieved reports of studies included in the index meta-analyses that the systematic reviewers cited. It is possible that other papers relating to the studies exist, which contain additional results that are compatible with the index meta-analysis (e.g. results of cohort studies or randomized trials at later time points may have been presented in other papers which were not cited by the systematic reviewers). For this reason, we may have underestimated the true extent of multiplicity of results within studies. We were unable to translate and interpret the data from four included non-English language studies however, their absence is unlikely to have modified the observed extent of multiplicity. Finally, we only searched PubMed and Epistemonikos to find all SRs and included SRs written in English, so generalisation of our study findings to non-indexed SRs and non-English language SRs is potentially limited.

### Implications of this research for practice

Our study, in common with previous research ([9, 12, 13]), demonstrates that multiplicity of effect estimates in primary studies is very common, and is therefore an issue that systematic reviwers should prepare for when designing their review. Doing so will have multiple benefits. It will reduce post-hoc decision making, and in doing-so, provide greater assurance to a reader that the results have not been ‘cherry-picked’ for inclusion in the meta-analyses. Furthermore, specification of eligibility criteria and decision rules is likely to lessen the data extraction effort, requiring less information to be extracted from each primary study.

Our results suggest that in nutrition research, specification of eligibility criteria and decision rules to select results from among multiple unadjusted or covariate-adjusted analyses and overlapping samples of participants and different subgroups are most important to pre-specify in SRs that include non-randomised studies. On the other hand, methods used to select results arising from both final values and change from baseline values, and multiple time points are most important to pre-specify in SRs that include randomized trials with continuous outcomes. Furthermore, in the SR, we recommend reporting any modifications to the specified rules, or any additions that were introduced to cover multiplicity scenarios that had not been anticipated when designing the review.

The Cochrane Handbook for Systematic Reviews of Interventions version 6.1 [67] provides updated guidance to systematic reviewers about how to group interventions with multiple components or co-interventions or how to select from multiple comparisons and handle multiplicity of outcomes when conducting meta-analyses. In addition, the recently updated “Preferred Reporting Items for Systematic reviews and Meta-Analyses (PRISMA) statement” [68] includes a new item (10a), which recommends authors “List and define all outcomes for which data were sought. Specify whether all results that were compatible with each outcome domain in each study were sought (e.g. for all measures, time points, analyses), and if not, the methods used to decide which results to collect”. Implementation of recommendations from these sources will allow readers to understand the result selection process.

## 5. Conclusion

Our study found that in systematic reviews examining the effects of foods and diet, that multiplicity of results in the included primary studies was very common. Yet, pre-specification of decision rules to select from multiple results was not common. Systematic reviewers are encouraged to consider methods for dealing with multiplicity when designing their reviews. Doing so will limit the potential for selective inclusion of results, thus providing greater assurance to readers as to the trustworthiness of the review.

## Data Availability

Data are available upon request from the corresponding author.

## DECLARATIONS

### Contributors

All authors declare to meet the ICMJE conditions for authorship. MJP and JEM conceived the project. MJP, JEM, LB, ZD, SM, and CMK contributed to the design of the project. MJP, ZD, SM, and CMK screened articles for inclusion. RK, MJP, LB, ZD, SM, CMK, and EK collected data. RK analysed the data. RK wrote the first draft of the manuscript, which was revised in conjunction with MJP and JEM. JEM drafted sections of the manuscript. All authors were involved in revising the article critically for important intellectual content. All authors approved the final version of the article. MJP is the guarantor of this work.

