## Supplementary for "Methods used to select results to include in meta-analyses of nutrition research: a meta-research study"

**Supplementary files**

**Table S1. Modifications from the ROBUST study protocol**

| Original plan | Revised plan | Reason for modification |
|---|---|---|
| We will seek a sample of SRs with meta-analysis meeting the following criteria:<br>• includes studies of people of any ages (i.e. infants, children, adolescents, adults or elderly people) and backgrounds in the generally healthy population, including pregnant and breastfeeding women and people with common diet-related risk factors such as being overweight or having high blood pressure | We will seek a sample of SRs with meta-analysis meeting the following criteria:<br>• includes studies that enrolled (a) people of any age or background who were generally healthy, (b) a mixture of generally healthy people and people with diet-related risk factors (e.g. overweight, high blood pressure) or a particular health condition (e.g. type II diabetes or cardiovascular disease), or (c) people with non-specified health status; | During screening of full text articles, we discovered that many otherwise eligible SRs did not state the health status of participants in included studies, or included a mix of participants who were healthy or those who had a particular clinical condition. After screening 1,500 abstracts and retrieving 149 full text articles, 44 SRs were considered to meet the eligibility criteria; however, on closer inspection, only 7 SRs included studies restricted to “healthy people”, 12 SRs included a mixture of studies of healthy people and people with a health condition (e.g. type II diabetes), and in 25 SRs the health status of participants was not clear (e.g. the authors said they included studies of “adults” and the characteristics of included studies presented provided no information on health status or risk factors). We have no reason to believe that SRs meeting the original inclusion criteria would be systematically different from SRs meeting the revised criteria. Therefore, rather than continue screening abstracts meeting the more stringent, original criteria, we decided to include SRs meeting the less stringent, revised criteria. |

| Original plan | Revised plan | Reason for modification |
|---|---|---|
| We will include meta-analyses of binary outcomes and meta-analyses of continuous outcomes. | We will include meta-analyses of non-continuous outcomes (e.g. binary, count, time-to-event) and meta-analyses of continuous outcomes. | During screening of articles we decided to expand eligibility from meta-analyses of “binary” to meta-analyses of “non-continuous” outcomes given the types of multiplicity affecting these meta-analyses are similar. |
| No plan for dealing with the scenario whereby screening of full text articles results in more than 25 SRs of one type (with binary or continuous outcomes). | If, during the screening process the total number of eligible SRs identified exceeded the target of 25 SRs with a meta-analysis of a non-continuous outcome and 25 SRs with a meta-analysis of a continuous outcome, we will randomly sample 25 SRs of each type. | By the time we had identified 25 eligible systematic reviews with meta-analysis of a continuous outcome, we had identified 74 eligible SRs with meta-analysis of a non-continuous outcome. Rather than include all SRs with meta-analysis of a non-continuous outcome in the study, we decided to randomly select 25 SRs. |
| Following piloting, two investigators will independently collect data from all remaining index meta-analyses and included studies. | Following piloting, two investigators will independently collect data on half of the remaining index meta-analyses and their included studies, while one investigator will collect data on the other half of the remaining index meta-analyses and their included studies. We will randomly select the SRs which will have their data collected by two investigators. | We decided to reduce the workload for investigators responsible for collecting data. |
| We will select a random sample of 50 SRs. From each SR, one investigator will select one meta-analysis of a binary or continuous outcome for assessment, stratifying the selection so that the final sample includes 25 binary and 25 continuous outcome meta-analyses. We will extract data from all SRs | We will select a random sample of 50 SRs, 25 of which will have a non-continuous index meta-analysis and 25 of which will have a continuous index meta-analysis. However, we will only extract data from SRs with less than 20 studies included in the index meta-analysis. | We decided to reduce the workload for investigators responsible for collecting data. |

| Original plan | Revised plan | Reason for modification |
|---|---|---|
| selected, regardless of the number of included studies in the index meta-analysis. | | |
| No plan for handling subgroup effect estimates in studies. | We will only extract subgroup effect estimates from studies if (a) they were the most compatible with the index meta-analysis (e.g. if the index meta-analysis is defined as "cancer mortality in women", we will only extract subgroup estimates of cancer mortality for women); (b) they were the only effect estimates available in a paper and both were compatible with the index meta-analysis (e.g. if the index meta-analysis is defined as "cancer mortality" and study investigators report an estimate for men and an estimate for women, but not the combined estimate, we will extract both subgroup estimates); or (c) the systematic reviewers included subgroup effect estimates in the index meta-analysis. We will not extract subgroup effect estimates from studies if none of the 3 criteria above are met, and if an estimate based on the total sample of participants was available in the paper. | Subgroup effect estimates are less precise and less generalisable than effect estimates based on the total sample. For this reason, selecting an effect estimate based on the total sample (when available) ahead of the subgroup effect estimates should not be considered selective inclusion of results by reviewers. |

### Supplement 2:

### Search Strategies

**PubMed search**

1. Nutritional Sciences[mh] OR Nutritional Physiological Phenomena[mh] OR Nutrition Assessment[mh] OR Nutrition Therapy[mh] OR Nutrition Policy[mh] OR Nutritional and Metabolic Diseases[mh] OR nutrition*[tiab] OR diet[tiab] OR feeding[tiab] OR dietary[tiab] OR breastfeed*[tiab] OR breast feed*[tiab] OR lactation[tiab] OR bottle feed*[tiab] OR complementary feeding[tiab] OR weaning[tiab] OR enteral[tiab] OR parenteral[tiab] OR Feeding Methods[mh] OR nutritional status[tiab] OR overweight[tiab] OR obese[tiab] OR obesity[tiab] OR overnutrition[tiab] OR over nutrition[tiab] OR undernourished[tiab] OR overnourished[tiab] OR wasted[tiab] OR wasting[tiab] OR stunting[tiab] OR stunted[tiab] OR underweight[tiab] OR undernutrition[tiab] OR under nutrition[tiab] OR body weight[tiab] OR anthropometry[tiab] OR Body Weights and Measures[mh] OR growth monitoring[tiab] OR food[tiab] OR food labelling[mh] OR food assistance[mh] OR supplementary feeding[tiab] OR diet therapy[mh] OR food and beverages[mh] OR vegetable*[tiab] OR fruit*[tiab] OR meat[tiab] OR dairy[tiab] OR dietary fat*[tiab] OR starch*[tiab] OR cereal[tiab] OR food-drug interactions[mh] OR food supply[mh] OR feeding behavio*[tiab] OR eating behavio*[tiab] OR food pattern*[tiab] OR food hypersensitivity[mh] OR food deprivation[mh] OR food, organic [mh] OR micronutrient*[tiab] OR vitamin*[tiab] OR thiamin[tiab] OR riboflavin[tiab] OR niacin[tiab] OR pantothenic acid[tiab] OR pyridoxine[tiab] OR pyridoxal[tiab] OR pyridoxamine[tiab] OR biotin[tiab] OR folic acid[tiab] OR folate[tiab]OR cyanocobalamin[tiab] OR choline[tiab] OR retinol[tiab] OR ascorbic acid[tiab] OR tocopherol[tiab] OR carotenoids[tiab] OR carotene[tiab] OR cryptoxanthin[tiab] OR lutein[tiab] OR lycopene[tiab] OR zeaxanthin[tiab] OR minerals[tiab] OR calcium[tiab] OR chloride[tiab] OR magnesium[tiab] OR phosphorus[tiab] OR potassium[tiab] OR sodium[tiab] OR iron[tiab] OR sulphur[tiab] OR trace element*[tiab] OR boron[tiab] OR cobalt[tiab] OR chromium[tiab] OR copper[tiab] OR fluoride[tiab] OR iodine[tiab] OR iron[tiab] OR manganese[tiab] OR molybdenum[tiab] OR selenium[tiab] OR zinc[tiab] OR trace metal*[tiab] OR macronutrient*[tiab] OR carbohydrate*[tiab] OR dietary protein*[tiab] OR saturated fat*[tiab]
OR unsaturated fat*[tiab] OR mono unsaturated fat*[tiab] OR monounsaturated fat*[tiab] OR poly unsaturated fat*[tiab] OR polyunsaturated fat*[tiab] OR trans fat*[tiab] OR dietary

fibre[tiab] OR dietary fiber[tiab] OR dietary salt[tiab] OR table salt[tiab] OR soft drink[tiab] OR fruit juice[tiab] OR vegetable juice[tiab] OR milk[tiab] OR tea[tiab] OR coffee[tiab] OR energy drink*[tiab] OR carbonated beverage*[tiab] OR carbonated drink*[tiab] OR prebiotics[tiab] OR probiotics[tiab] OR glycemic load[tiab] OR glycemic index[tiab] OR glycaemic load[tiab] OR glycaemic index[tiab] OR calories[tiab] OR kilocalories[tiab] OR kilojoules[tiab] OR caloric intake[tiab] OR energy intake[tiab]
2. meta-analysis[pt] OR meta-analy*[ti]
3. ("2018/01/01"[PDAT] : "2019/06/30"[PDAT])
4. 1 AND 2 AND 3

**Epistemonikos search**
1. (advanced_title_en:(nutri* OR diet*) OR advanced_abstract_en:(nutri* OR diet*)) [Filters: protocol=no, min_year=2018, max_year=2019]

**Supplementary Table S2: Data collection form**

| | # | Variable / Field Name | Field Label<br>*Field Note* | Field Attributes (Field Type, Validation, Choices, Calculations, etc.) |
|---|---|---|---|---|
| Instrument: **SR protocol characteristics** (sr_protocol_characteristics) | | | | |
| | 1 | record_id | Record ID: | text |
| | 2 | sr_id | SR ID (stated on page 1 of PDF): | text |
| | 3 | initials | Enter initials of data collector: | text |
| | 4 | index_ma | Specify the index meta-analysis outcome (stated on pg 1 of the PDF): | text |
| | 5 | protocol_avail | Is a protocol or registration record available for the SR? | radio<br>1 Both a protocol and registration record are available<br>2 Only a protocol is available<br>3 Only a registration record is available<br>4 Neither are available |
| | 6 | srp_year<br>Show the field ONLY if:<br>[protocol_avail] = '1' or [protocol_avail] = '2' | Specify the year of publication of the protocol: | text |
| | 7 | regist_year<br>Show the field ONLY if: | Specify the year the SR was first registered: | text |

| | | | | |
|---|---|---|---|---|
| | | [protocol_avail] = '1' or [protocol_avail] = '3' | | |
| | 8 | srp_e<br>Show the field ONLY if:<br>[protocol_avail] = '1' or [protocol_avail] = '2' or [protocol_avail] = '3' | Section Header: *Eligibility criteria and decision rules*<br>Review authors pre-specified...(select ALL of the following that apply): | checkbox<br>1 \| srp_e___1 \| ...eligible measurement instruments for [index_ma]<br>2 \| srp_e___2 \| ...eligible definitions/diagnostic criteria for [index_ma]<br>3 \| srp_e___3 \| ...eligible cut-points on an instrument for [index_ma]<br>4 \| srp_e___4 \| ...eligible time points for [index_ma]<br>5 \| srp_e___5 \| ...eligible interventions/exposures<br>6 \| srp_e___6 \| ...whether final values or change from baseline values (or both) for [index_ma] were eligible<br>7 \| srp_e___7 \| ...which analysis samples for [index_ma] were eligible (e.g. intention-to-treat, per-protocol or as-treated)<br>8 \| srp_e___8 \| ...whether unadjusted or covariate-adjusted analyses (or both) for [index_ma] were eligible<br>9 \| srp_e___9 \| ...whether period or paired analyses (or both) for [index_ma] were eligible (from crossover trials)<br>10 \| srp_e___10 \| ...which information sources were eligible (e.g. journal article, trials results register) |
| | 9 | srp_dr<br>Show the field ONLY if:<br>[protocol_avail] = '1' or [protocol_avail] = '2' or [protocol_avail] = '3' | Review authors pre-specified...(select ALL of the following that apply): | checkbox<br>1 \| srp_dr___1 \| ...a decision rule to handle results arising from multiple measurement instruments for [index_ma]<br>2 \| srp_dr___2 \| ...a decision rule to handle results arising from multiple definitions/diagnostic criteria for [index_ma] |

| | | | | | | |
|---|---|---|---|---|---|---|
| | | | | 3 | srp_dr___3 | ...a decision rule to handle results arising from multiple cut-points on an instrument for [index_ma] |
| | | | | 4 | srp_dr___4 | ...a decision rule to handle results arising from multiple time points for [index_ma] |
| | | | | 5 | srp_dr___5 | ...a decision rule to handle results arising from multiple interventions/exposures |
| | | | | 6 | srp_dr___6 | ...a decision rule to handle results arising from both final values and change from baseline values for [index_ma] |
| | | | | 7 | srp_dr___7 | ...a decision rule to handle results arising from multiple analysis samples (e.g. intention-to-treat, per-protocol or as-treated) for [index_ma] |
| | | | | 8 | srp_dr___8 | ...a decision rule to handle results arising from multiple unadjusted or covariate-adjusted analyses for [index_ma] |
| | | | | 9 | srp_dr___9 | ...a decision rule to handle results arising from period and paired analyses for [index_ma] in crossover trials |
| | | | | 10 | srp_dr___10 | ...a decision rule to handle results arising from multiple information sources (e.g. journal article, trials results register) |
| | | | | 11 | srp_dr___11 | ...a decision rule to handle results arising from overlapping samples of participants (e.g. subgroup of men of all ages and subgroup of older adults which includes men) |
| | | | | 12 | srp_dr___12 | ...a decision rule to handle results arising from another source of multiplicity |

| | | | | |
|---|---|---|---|---|
| | 10 | srp_e_scale_txt<br>Show the field ONLY if:<br>[srp_e(1)] = '1' | Specify the eligible measurement instruments for [index_ma]: | notes |
| | 11 | srp_dr_scale_txt<br>Show the field ONLY if:<br>[srp_dr(1)] = '1' | Specify the decision rule to handle results arising from multiple measurement instruments for [index_ma]: | notes |
| | 12 | srp_e_event_txt<br>Show the field ONLY if:<br>[srp_e(2)] = '1' | Specify the eligible definitions/diagnostic criteria for [index_ma]: | notes |
| | 13 | srp_dr_event_txt<br>Show the field ONLY if:<br>[srp_dr(2)] = '1' | Specify the decision rule to handle results arising from multiple definitions/diagnostic criteria for [index_ma]: | notes |
| | 14 | srp_e_cutpoint_txt<br>Show the field ONLY if:<br>[srp_e(3)] = '1' | Specify the eligible cut-points on an instrument for [index_ma]: | notes |
| | 15 | srp_dr_cutpoint_txt<br>Show the field ONLY if:<br>[srp_dr(3)] = '1' | Specify the decision rule to handle results arising from multiple cut-points on an instrument for [index_ma]: | notes |

| | | | | |
|---|---|---|---|---|
| | 16 | srp_e_time_txt<br>Show the field ONLY if:<br>[srp_e(4)] = '1' | Specify the eligible time points for [index_ma]: | notes |
| | 17 | srp_dr_time_txt<br>Show the field ONLY if:<br>[srp_dr(4)] = '1' | Specify the decision rule to handle results arising from multiple time points for [index_ma]: | notes |
| | 18 | srp_e_group_txt<br>Show the field ONLY if:<br>[srp_e(5)] = '1' | Specify the eligible interventions/exposures (incl. comparators): | notes |
| | 19 | srp_dr_group_txt<br>Show the field ONLY if:<br>[srp_dr(5)] = '1' | Specify the decision rule to handle results arising from multiple interventions/exposures: | notes |
| | 20 | srp_e_fvcs_txt<br>Show the field ONLY if:<br>[srp_e(6)] = '1' | Specify which values (i.e. final, change from baseline or both) for [index_ma] were eligible: | notes |
| | 21 | srp_dr_fvcs_txt<br>Show the field ONLY if:<br>[srp_dr(6)] = '1' | Specify the decision rule to handle results arising from both final and change from baseline values for [index_ma]: | notes |
| | 22 | srp_e_itt_txt<br>Show the field ONLY if:<br>[srp_e(7)] = '1' | Specify which analysis samples (e.g. intention-to-treat, per-protocol) | notes |

| | | | | |
|---|---|---|---|---|
| | | | for [index_ma] were eligible: | |
| | 23 | srp_dr_itt_txt<br>Show the field ONLY if:<br>[srp_dr(7)] = '1' | Specify the decision rule to handle results arising from multiple analysis samples (e.g. intention-to-treat, per-protocol) for [index_ma]: | notes |
| | 24 | srp_e_unadj_txt<br>Show the field ONLY if:<br>[srp_e(8)] = '1' | Specify which unadjusted and covariate-adjusted analyses for [index_ma] were eligible: | notes |
| | 25 | srp_dr_unadj_txt<br>Show the field ONLY if:<br>[srp_dr(8)] = '1' | Specify the decision rule to handle results arising from multiple unadjusted and covariate-adjusted analyses for [index_ma]: | notes |
| | 26 | srp_e_cross_txt<br>Show the field ONLY if:<br>[srp_e(9)] = '1' | Specify which analyses in crossover trials (e.g. first period, second period, paired) were eligible: | notes |
| | 27 | srp_dr_cross_txt<br>Show the field ONLY if:<br>[srp_dr(9)] = '1' | Specify the decision rule to handle results arising from period and | notes |

| | | | | |
|---|---|---|---|---|
| | | | paired analyses in crossover trials: | |
| | 28 | srp_e_source_txt<br>Show the field ONLY if:<br>[srp_e(10)] = '1' | Specify which information sources (e.g. journal article, trials results register) were eligible: | notes |
| | 29 | srp_dr_source_txt<br>Show the field ONLY if:<br>[srp_dr(10)] = '1' | Specify the decision rule to handle results arising from multiple information sources (e.g. journal article, trials results register): | notes |
| | 30 | srp_dr_overlap_txt<br>Show the field ONLY if:<br>[srp_dr(11)] = '1' | Specify the decision rule to handle results arising from overlapping samples of participants: | notes |
| | 31 | srp_dr_other_txt<br>Show the field ONLY if:<br>[srp_dr(12)] = '1' | Specify the decision rule(s) to handle results arising from other sources of multiplicity for [index_ma]: | notes |
| | 32 | srp_notes | Notes | notes |
| | 33 | sr_protocol_characteristics_complete | Section Header: *Form Status*<br>Complete? | dropdown<br>0 Incomplete<br>1 Unverified |

| | | | | |
|---|---|---|---|---|
| | | | | 2 Complete |
| Instrument: **SR characteristics** (sr_characteristics) | | | | |
| | 34 | sr_title | Specify the title of the systematic review: | text |
| | 35 | sr_journal | Specify the journal: | text |
| | 36 | sr_year | Specify the year of publication: | text |
| | 37 | sr_country | Specify the country of the corresponding author of the SR: | text |
| | 38 | sr_funding | What was the source of funding for the SR? | radio<br>1 Non-profit<br>2 For-profit<br>3 Mixed<br>4 No funding<br>5 Not reported |
| | 39 | for_profit_org<br>Show the field ONLY if:<br>[sr_funding] = '2' or [sr_funding] = '3' | What type of for-profit organisation funded the SR? | radio<br>1 Food industry<br>2 Other industry |
| | 40 | for_profit_org_txt<br>Show the field ONLY if: | Record the name of the for-profit funder: | text |

| | | | | |
|---|---|---|---|---|
| | | [sr_funding] = '2' or [sr_funding] = '3' | | |
| | 41 | sr_coi | Did any of the systematic reviewers disclose financial conflicts of interest? | radio<br>1 Conflict of interest present (i.e. at least one systematic reviewer reported a financial conflict of interest of any type, excluding current study funding or industry employment)<br>2 No conflict of interest (i.e. all systematic reviewers stated they had no conflicts)<br>3 Missing (i.e. no disclosure statement) |
| | 42 | sr_coi_txt<br>Show the field ONLY if:<br>[sr_coi] = '1' | Record verbatim the conflict(s) of interest disclosed by the authors: | notes |
| | 43 | sr_affiliat | What type of affiliation did the corresponding author of the SR have? | radio<br>1 Food industry<br>4 Other industry<br>2 Non-industry<br>3 Mixed<br>5 Unclear |
| | 44 | ma_n_studies | Section Header: *General characteristics of the index meta-analysis (i.e. [index_ma])* | text |

| | | | | |
|---|---|---|---|---|
| | | | Specify the total number of studies included in the index meta-analysis (stated on page 1 of PDF): | |
| | 45 | ma_n_patients | Specify the total number of participants included in the index meta-analysis (sum values presented on the forest plot/table or type 'NR' if sample sizes were not available): | text |
| | 46 | ma_patients | Specify the type of participants included in studies included in the index meta-analysis (e.g. "generally healthy", "normal weight"): | text |
| | 47 | ma_intervention | Specify the intervention/exposure investigated in the index meta-analysis (e.g. "red meat 3 times a week"): | text |
| | 48 | ma_comparator | Specify the comparator investigated in the index meta-analysis | text |

| | | | | |
|---|---|---|---|---|
| | | | (e.g. "red meat once a month"): | |
| | 49 | ma_study_type | What type of studies were included in the index meta-analysis? | radio<br>1 Randomized trial<br>2 Non-randomized study<br>3 Both randomized trials and non-randomized studies |
| | 50 | ma_study_type_spec | Indicate the designs of the studies included in the index meta-analysis as specified by the systematic reviewers (e.g. cohort study, case-control study): | text |
| | 51 | ma_outcome_spec | Record how the index meta-analysis was specified (e.g. "weight" or "weight at 6 weeks" or "change in weight at 6 weeks"): | text |
| | 52 | ma_domain | Specify the outcome domain investigated in the index meta-analysis (e.g. all-cause mortality, body weight): | text |
| | 53 | ma_outcome_label | Specify the label given by the review authors to the index meta-analysis outcome: | radio<br>1 Primary<br>2 Secondary |

| | | | | |
|---|---|---|---|---|
| | | | | 3 Unlabelled |
| | 54 | ma_model | Specify the model for the index meta-analysis: | radio<br>1 Fixed-effect<br>2 Random-effects<br>3 Unclear |
| | 55 | sr_e | Section Header: *Eligibility criteria and decision rules*<br>Review authors reported...(select ALL of the following that apply): | checkbox<br>1 sr_e___1 ...eligible measurement instruments for [index_ma]<br>2 sr_e___2 ...eligible definitions/diagnostic criteria for [index_ma]<br>3 sr_e___3 ...eligible cut-points on an instrument for [index_ma]<br>4 sr_e___4 ...eligible time points for [index_ma]<br>5 sr_e___5 ...eligible interventions/exposures<br>6 sr_e___6 ...whether final values or change from baseline values (or both) for [index_ma] were eligible<br>7 sr_e___7 ...which analysis samples for [index_ma] were eligible (e.g. intention-to-treat, per-protocol or as-treated)<br>8 sr_e___8 ...whether unadjusted or covariate-adjusted analyses (or both) for [index_ma] were eligible<br>9 sr_e___9 ...whether period or paired analyses (or both) for [index_ma] were eligible (from crossover trials)<br>10 sr_e___10 ...which information sources were eligible (e.g. journal article, trials results register) |
| | 56 | sr_dr | Review authors reported...(select ALL | checkbox |

<table>
<tr>
<td></td>
<td></td>
<td></td>
<td>of the following that apply):</td>
<td>
<table>
<tr><td>1</td><td>sr_dr___1</td><td>...a decision rule to handle results arising from multiple measurement instruments for [index_ma]</td></tr>
<tr><td>2</td><td>sr_dr___2</td><td>...a decision rule to handle results arising from multiple definitions/diagnostic criteria for [index_ma]</td></tr>
<tr><td>3</td><td>sr_dr___3</td><td>...a decision rule to handle results arising from multiple cut-points on an instrument for [index_ma]</td></tr>
<tr><td>4</td><td>sr_dr___4</td><td>...a decision rule to handle results arising from multiple time points for [index_ma]</td></tr>
<tr><td>5</td><td>sr_dr___5</td><td>...a decision rule to handle results arising from multiple interventions/exposures</td></tr>
<tr><td>6</td><td>sr_dr___6</td><td>...a decision rule to handle results arising from both final values and change from baseline values for [index_ma]</td></tr>
<tr><td>7</td><td>sr_dr___7</td><td>...a decision rule to handle results arising from multiple analysis samples (e.g. intention-to-treat, per-protocol or as-treated) for [index_ma]</td></tr>
<tr><td>8</td><td>sr_dr___8</td><td>...a decision rule to handle results arising from multiple unadjusted or covariate-adjusted analyses for [index_ma]</td></tr>
<tr><td>9</td><td>sr_dr___9</td><td>...a decision rule to handle results arising from period and paired analyses for [index_ma] in crossover trials</td></tr>
<tr><td>10</td><td>sr_dr___10</td><td>...a decision rule to handle results arising from multiple information sources (e.g. journal article, trials results register)</td></tr>
<tr><td>11</td><td>sr_dr___11</td><td>...a decision rule to handle results arising from overlapping samples of participants (e.g. subgroup of men of all ages and subgroup of older adults which includes men)</td></tr>
</table>
</td>
</tr>
</table>

| | | | | |
|---|---|---|---|---|
| | | | | 12 \| sr_dr___12 \| ...a decision rule to handle results arising from another source of multiplicity |
| | 57 | sr_e_scale_txt<br>Show the field ONLY if:<br>[sr_e(1)] = '1' | Specify the eligible measurement instruments for [index_ma]: | notes |
| | 58 | sr_dr_scale_txt<br>Show the field ONLY if:<br>[sr_dr(1)] = '1' | Specify the decision rule to handle results arising from multiple measurement instruments for [index_ma]: | notes |
| | 59 | sr_e_event_txt<br>Show the field ONLY if:<br>[sr_e(2)] = '1' | Specify the eligible definitions/diagnostic criteria for [index_ma]: | notes |
| | 60 | sr_dr_event_txt<br>Show the field ONLY if:<br>[sr_dr(2)] = '1' | Specify the decision rule to handle results arising from multiple definitions/diagnostic criteria for [index_ma]: | notes |
| | 61 | sr_e_cutpoint_txt<br>Show the field ONLY if:<br>[sr_e(3)] = '1' | Specify the eligible cut-points on an instrument for [index_ma]: | notes |
| | 62 | sr_dr_cutpoint_txt<br>Show the field ONLY if:<br>[sr_dr(3)] = '1' | Specify the decision rule to handle results arising from multiple cut-points on an | notes |

| | | | | |
|---|---|---|---|---|
| | | | instrument for [index_ma]: | |
| | 63 | sr_e_time_txt<br>Show the field ONLY if:<br>[sr_e(4)] = '1' | Specify the eligible time points for [index_ma]: | notes |
| | 64 | sr_dr_time_txt<br>Show the field ONLY if:<br>[sr_dr(4)] = '1' | Specify the decision rule to handle results arising from multiple time points for [index_ma]: | notes |
| | 65 | sr_e_group_txt<br>Show the field ONLY if:<br>[sr_e(5)] = '1' | Specify the eligible interventions/exposures (incl. comparators): | notes |
| | 66 | sr_dr_group_txt<br>Show the field ONLY if:<br>[sr_dr(5)] = '1' | Specify the decision rule to handle results arising from multiple interventions/exposures: | notes |
| | 67 | sr_e_fvcs_txt<br>Show the field ONLY if:<br>[sr_e(6)] = '1' | Specify which values (i.e. final, change from baseline or both) for [index_ma] were eligible: | notes |
| | 68 | sr_dr_fvcs_txt<br>Show the field ONLY if:<br>[sr_dr(6)] = '1' | Specify the decision rule to handle results arising from both final and change from baseline values for [index_ma]: | notes |

| | | | | |
|---|---|---|---|---|
| | 69 | sr_e_itt_txt<br>Show the field ONLY if:<br>[sr_e(7)] = '1' | Specify which analysis samples (e.g. intention-to-treat, per-protocol) for [index_ma] were eligible: | notes |
| | 70 | sr_dr_itt_txt<br>Show the field ONLY if:<br>[sr_dr(7)] = '1' | Specify the decision rule to handle results arising from multiple analysis samples (e.g. intention-to-treat, per-protocol) for [index_ma]: | notes |
| | 71 | sr_e_unadj_txt<br>Show the field ONLY if:<br>[sr_e(8)] = '1' | Specify which unadjusted and covariate-adjusted analyses for [index_ma] were eligible: | notes |
| | 72 | sr_dr_unadj_txt<br>Show the field ONLY if:<br>[sr_dr(8)] = '1' | Specify the decision rule to handle results arising from multiple unadjusted and covariate-adjusted analyses for [index_ma]: | notes |
| | 73 | sr_e_cross_txt<br>Show the field ONLY if:<br>[sr_e(9)] = '1' | Specify which analyses in crossover trials (e.g. first period, second period, paired) were eligible: | notes |

| | | | | |
|---|---|---|---|---|
| | 74 | sr_dr_cross_txt<br>Show the field ONLY if:<br>[sr_dr(9)] = '1' | Specify the decision rule to handle results arising from period and paired analyses in crossover trials: | notes |
| | 75 | sr_e_source_txt<br>Show the field ONLY if:<br>[sr_e(10)] = '1' | Specify which information sources (e.g. journal article, trials results register) were eligible: | notes |
| | 76 | sr_dr_source_txt<br>Show the field ONLY if:<br>[sr_dr(10)] = '1' | Specify the decision rule to handle results arising from multiple information sources (e.g. journal article, trials results register): | notes |
| | 77 | sr_dr_overlap_txt<br>Show the field ONLY if:<br>[sr_dr(11)] = '1' | Specify the decision rule to handle results arising from overlapping samples of participants: | notes |
| | 78 | sr_dr_other_txt<br>Show the field ONLY if:<br>[sr_dr(12)] = '1' | Specify the decision rule(s) to handle results arising from other sources of multiplicity for [index_ma]: | notes |
| | 79 | sr_notes | Notes | notes |

| | | | | |
|---|---|---|---|---|
| | 80 | sr_characteristics_complete | Section Header: *Form Status*<br>Complete? | dropdown<br>0 Incomplete<br>1 Unverified<br>2 Complete |
| Instrument: **Meta-analysis study data** (metaanalysis_study_data) | | | | |
| | 81 | ma_study_id | Study ID (record the ID appearing on the forest plot/table for the index meta-analysis): | text |
| | 82 | ma_st_measure | Specify the study effect measure for [index_ma]: | radio<br>1 Risk ratio<br>2 Odds ratio<br>3 Hazard ratio<br>4 Mean difference<br>5 Standardized mean difference |
| | 83 | ma_st_event1<br>Show the field ONLY if:<br>[ma_st_measure] = '1' or [ma_st_measure] = '2' or [ma_st_measure] = '3' | Number of events in intervention/exposure group (type 'NR' if not reported): | text |
| | 84 | ma_st_total1<br>Show the field ONLY if:<br>[ma_st_measure] = '1' or [ma_st_measure] = '2' or [ma_st_measure] = '3' | Sample size of intervention/exposure group (type 'NR' if not reported): | text |

| | | | | |
|---|---|---|---|---|
| | 85 | ma_st_event2<br>Show the field ONLY if:<br>[ma_st_measure] = '1' or [ma_st_measure] = '2' or [ma_st_measure] = '3' | Number of events in comparator group (type 'NR' if not reported): | text |
| | 86 | ma_st_total2<br>Show the field ONLY if:<br>[ma_st_measure] = '1' or [ma_st_measure] = '2' or [ma_st_measure] = '3' | Sample size of comparator group (type 'NR' if not reported): | text |
| | 87 | ma_st_mean1<br>Show the field ONLY if:<br>[ma_st_measure] = '4' or [ma_st_measure] = '5' | Mean of intervention/exposure group (type 'NR' if not reported): | text |
| | 88 | ma_st_sd1<br>Show the field ONLY if:<br>[ma_st_measure] = '4' or [ma_st_measure] = '5' | Standard deviation of intervention/exposure group (type 'NR' if not reported): | text |
| | 89 | ma_st_n1<br>Show the field ONLY if:<br>[ma_st_measure] = '4' or [ma_st_measure] = '5' | Sample size of intervention/exposure group (type 'NR' if not reported): | text |
| | 90 | ma_st_mean2<br>Show the field ONLY if:<br>[ma_st_measure] = '4' or [ma_st_measure] = '5' | Mean of comparator group (type 'NR' if not reported): | text |
| | 91 | ma_st_sd2<br>Show the field ONLY if: | Standard deviation of comparator group (type 'NR' if not reported): | text |

| | | | | |
|---|---|---|---|---|
| | | [ma_st_measure] = '4' or [ma_st_measure] = '5' | | |
| | 92 | ma_st_n2<br>Show the field ONLY if:<br>[ma_st_measure] = '4' or [ma_st_measure] = '5' | Sample size of comparator group (type 'NR' if not reported): | text |
| | 93 | ma_st_est | Study effect estimate (type 'NR' if not reported): | text |
| | 94 | ma_st_seest | Standard error of the study effect estimate (type 'NR' if not reported): | text |
| | 95 | ma_st_lciest | Lower 95% confidence interval of the study effect estimate (type 'NR' if not reported): | text |
| | 96 | ma_st_uciest | Upper 95% confidence interval of the study effect estimate (type 'NR' if not reported): | text |
| | 97 | ma_st_ci_level | If the confidence interval (CI) was not a 95% CI, specify the CI level (e.g. 90% CI, 99% CI); otherwise leave blank | text |

| | | | | |
|---|---|---|---|---|
| | 98 | ma_st_direction | Specify the direction of the study effect estimate (note: ignore the statistical significance of the effect, just focus on the direction): | radio<br>1 Favours intervention or exposure<br>2 Favours comparator<br>3 Neutral (e.g. RR is somewhere between 0.95 and 1.05 or MD is somewhere between -0.05 and 0.05)<br>4 Unclear |
| | 99 | ma_st_retrieve | Did the review authors state that data for this particular result were retrieved from study authors? | yesno<br>1 Yes<br>0 No |
| | 100 | ma_st_manip | Did the review authors state that data for this particular result required algebraic transformation to include in meta-analysis (e.g. convert SE to SD)? | yesno<br>1 Yes<br>0 No |
| | 101 | ma_st_transl | Did the review authors state that data for this particular result originated from a report translated into English? | yesno<br>1 Yes<br>0 No |
| | 102 | ma_st_impute | Did the review authors state that data for this particular result was included in the meta- | yesno<br>1 Yes<br>0 No |

| | | | | |
|---|---|---|---|---|
| | | | analysis by using a method of imputation? | |
| | 103 | ma_st_impute_txt<br>Show the field ONLY if:<br>[ma_st_impute] = '1' | Specify the method of imputation: | text |
| | 104 | ma_st_notes | Notes | notes |
| | 105 | metaanalysis_study_data_complete | Section Header: *Form Status*<br>Complete? | dropdown<br>0 Incomplete<br>1 Unverified<br>2 Complete |
| Instrument: **Meta-analytic effect** (metaanalytic_effect) | | | | |
| | 106 | ma_measure | Specify the meta-analytic effect measure for [index_ma]: | radio<br>1 Risk ratio<br>2 Odds ratio<br>3 Hazard ratio<br>4 Mean difference<br>5 Standardized mean difference |
| | 107 | ma_est | Meta-analytic effect estimate: | text |
| | 108 | ma_seest | Standard error of the meta-analytic effect estimate (type 'NR' if not reported): | text |

| | | | |
|---|---|---|---|
| 109 | ma_lciest | Lower 95% confidence interval of the meta-analytic effect estimate (type 'NR' if not reported): | text |
| 110 | ma_uciest | Upper 95% confidence interval of the meta-analytic effect estimate (type 'NR' if not reported): | text |
| 111 | ma_ci_level | If the confidence interval (CI) was not a 95% CI, specify the CI level (e.g. 90% CI, 99% CI); otherwise leave blank | text |
| 112 | ma_direction | Specify the direction of the meta-analytic effect estimate (note: ignore the statistical significance of the effect, just focus on the direction): | radio<br>1 Favours intervention or exposure<br>2 Favours comparator<br>3 Neutral (e.g. RR is somewhere between 0.95 and 1.05 or MD is somewhere between -0.05 and 0.05)<br>4 Unclear |
| 113 | ma_location | Specify the location of the meta-analysis in the article (e.g. Figure 1, Supp Table 2): | text |

| | | | | |
|---|---|---|---|---|
| | 114 | ma_conclusion | What type of conclusion did the authors draw about the intervention/exposure with regards to [index_ma]? | radio<br>1 Favourable (e.g. stated that the food/diet had a beneficial/positive effect on [index_ma])<br>2 Unfavourable (e.g. stated that the food/diet had a harmful/negative effect on [index_ma])<br>3 Neutral (e.g. stated that the food/diet had neither a beneficial or harmful effect on [index_ma])<br>4 Unclear<br>5 No conclusion about [index_ma] drawn |
| | 115 | ma_conclusion_ab_txt<br>Show the field ONLY if:<br>[ma_conclusion] = '1' or [ma_conclusion] = '2' or [ma_conclusion] = '3' or [ma_conclusion] = '5' | Extract verbatim the conclusion reported in the abstract: | notes |
| | 116 | ma_conclusion_txt<br>Show the field ONLY if:<br>[ma_conclusion] = '1' or [ma_conclusion] = '2' or [ma_conclusion] = '3' or [ma_conclusion] = '4' | Extract verbatim the conclusion reported in the main text: | notes |
| | 117 | ma_notes | Notes | notes |
| | 118 | metaanalytic_effect_complete | Section Header: *Form Status*<br>Complete? | dropdown<br>0 Incomplete<br>1 Unverified |

| | | | | |
|---|---|---|---|---|
| | | | | 2 Complete |
| Instrument: **Study data**(study_data) | | | | |
| | 119 | study_id | Study ID (record the ID in the filename of the PDF): | text |
| | 120 | st_data_avail | Was there enough data available in the paper to extract or calculate a result compatible with the index meta-analysis ([index_ma])? | yesno<br>1 Yes<br>0 No |
| | 121 | st_design<br>Show the field ONLY if:<br>[st_data_avail] = '1' | Indicate the design of the study (e.g. parallel-group randomized trial, crossover trial, cohort study, case-control study): | text |
| | 122 | st_measure<br>Show the field ONLY if:<br>[st_data_avail] = '1' | Specify the study effect measure for a result compatible with the index meta-analysis ([index_ma]): | radio<br>0 Risk ratio<br>1 Odds ratio<br>2 Hazard ratio<br>3 Rate ratio<br>4 Prevalence ratio<br>5 Mean difference<br>6 Standardized mean difference |

| | | | | |
|---|---|---|---|---|
| | 123 | st_def_event<br>Show the field ONLY if:<br>[st_measure] = '0' or [st_measure] = '1' or [st_measure] = '2' or [st_measure] = '3' or [st_measure] = '4' | Specify the definition/diagnostic criteria for the event (type 'NR' if not reported): | text |
| | 124 | st_scale<br>Show the field ONLY if:<br>[st_measure] = '5' or [st_measure] = '6' | Specify the outcome measurement instrument used to generate the result (type 'NR' if not reported): | text |
| | 125 | st_intervention<br>Show the field ONLY if:<br>[st_data_avail] = '1' | Specify the intervention/exposure (e.g. 'red meat 3 times a week'): | text |
| | 126 | st_comparator<br>Show the field ONLY if:<br>[st_data_avail] = '1' | Specify the comparator (e.g. 'red meat once a month'): | text |
| | 127 | st_time<br>Show the field ONLY if:<br>[st_data_avail] = '1' | Specify the time point for the result (e.g. 6 weeks; type 'NR' if not reported): | text |
| | 128 | st_itt<br>Show the field ONLY if:<br>[st_data_avail] = '1' | Specify the analysis sample for the result (type 'NR' if not reported): | radio<br>1 Intention-to-treat (ITT)<br>2 Modified ITT<br>3 Per-protocol<br>4 As-treated |

| | | | | |
|---|---|---|---|---|
| | | | | 5 Other<br>6 Unclear |
| | 129 | st_unadj<br>Show the field ONLY if:<br>[st_data_avail] = '1' | Was the result unadjusted or adjusted for covariates? | radio<br>1 Unadjusted<br>2 Adjusted<br>3 Unclear |
| | 130 | st_covar<br>Show the field ONLY if:<br>[st_unadj] = '2' | Which covariates were included in the model? | text |
| | 131 | st_other_id<br>Show the field ONLY if:<br>[st_data_avail] = '1' | Specify any other identifying characteristic of the result, e.g. 'subgroup analysis of men only' (type 'NA' if not applicable): | text |
| | 132 | st_event1<br>Show the field ONLY if:<br>[st_measure] = '0' or [st_measure] = '1' or [st_measure] = '2' or [st_measure] = '3' or [st_measure] = '4' | Number of events in intervention/exposure group (type 'NR' if not reported): | text |
| | 133 | st_total1<br>Show the field ONLY if:<br>[st_measure] = '0' or [st_measure] = '1' or [st_measure] = '2' | Sample size of intervention/exposure group (type 'NR' if not reported): | text |

| | | | | |
|---|---|---|---|---|
| | | or [st_measure] = '3' or [st_m easure] = '4' | | |
| | 134 | st_event2<br>Show the field ONLY if:<br>[st_measure] = '0' or [st_meas ure] = '1' or [st_measure] = '2' or [st_measure] = '3' or [st_m easure] = '4' | Number of events in comparator group (type 'NR' if not reported): | text |
| | 135 | st_total2<br>Show the field ONLY if:<br>[st_measure] = '0' or [st_meas ure] = '1' or [st_measure] = '2' or [st_measure] = '3' or [st_m easure] = '4' | Sample size of comparator group (type 'NR' if not reported): | text |
| | 136 | st_fvcs<br>Show the field ONLY if:<br>[st_measure] = '5' or [st_meas ure] = '6' | Which values were used to calculate means? | radio<br>1 Final values<br>2 Change from baseline values<br>3 Unclear |
| | 137 | st_meansd<br>Show the field ONLY if:<br>[st_measure] = '5' or [st_meas ure] = '6' | Were means and standard deviations reported for each group? | yesno<br>1 Yes<br>0 No |
| | 138 | st_mean1<br>Show the field ONLY if:<br>[st_measure] = '5' or [st_meas ure] = '6' | Mean of intervention/exposure group (type 'NR' if not reported): | text |

| | | | | |
|---|---|---|---|---|
| | 139 | st_sd1<br>Show the field ONLY if:<br>[st_measure] = '5' or [st_measure] = '6' | Standard deviation of intervention/exposure group (type 'NR' if not reported): | text |
| | 140 | st_n1<br>Show the field ONLY if:<br>[st_measure] = '5' or [st_measure] = '6' | Sample size of intervention/exposure group (type 'NR' if not reported or 'Paired N' if this was a before-after comparison): | text |
| | 141 | st_mean2<br>Show the field ONLY if:<br>[st_measure] = '5' or [st_measure] = '6' | Mean of comparator group (type 'NR' if not reported): | text |
| | 142 | st_sd2<br>Show the field ONLY if:<br>[st_measure] = '5' or [st_measure] = '6' | Standard deviation of comparator group (type 'NR' if not reported): | text |
| | 143 | st_n2<br>Show the field ONLY if:<br>[st_measure] = '5' or [st_measure] = '6' | Sample size of comparator group (type 'NR' if not reported or 'Paired N' if this was a before-after comparison): | text |
| | 144 | st_semean1<br>Show the field ONLY if:<br>[st_meansd] = '0' | Standard error of the mean of the intervention/exposure group (type 'NR' if not reported): | text |

| | | | | |
|---|---|---|---|---|
| | 145 | st_lcimean1<br>Show the field ONLY if:<br>[st_meansd] = '0' | Lower 95% confidence interval of the mean of the intervention/exposure group (type 'NR' if not reported): | text |
| | 146 | st_ucimean1<br>Show the field ONLY if:<br>[st_meansd] = '0' | Upper 95% confidence interval of the mean of the intervention/exposure group (type 'NR' if not reported): | text |
| | 147 | st_semean2<br>Show the field ONLY if:<br>[st_meansd] = '0' | Standard error of the mean of the comparator group (type 'NR' if not reported): | text |
| | 148 | st_lcimean2<br>Show the field ONLY if:<br>[st_meansd] = '0' | Lower 95% confidence interval of the mean of the comparator group (type 'NR' if not reported): | text |
| | 149 | st_ucimean2<br>Show the field ONLY if:<br>[st_meansd] = '0' | Upper 95% confidence interval of the mean of the comparator group (type 'NR' if not reported): | text |
| | 150 | st_est<br>Show the field ONLY if:<br>[st_data_avail] = '1' | Study effect estimate (type 'NR' if not reported): | text |

| | | | | |
|---|---|---|---|---|
| | 151 | st_seest<br>Show the field ONLY if:<br>[st_data_avail] = '1' | Standard error of the study effect estimate (type 'NR' if not reported): | text |
| | 152 | st_lciest<br>Show the field ONLY if:<br>[st_data_avail] = '1' | Lower 95% confidence interval of the study effect estimate (type 'NR' if not reported): | text |
| | 153 | st_uciest<br>Show the field ONLY if:<br>[st_data_avail] = '1' | Upper 95% confidence interval of the study effect estimate (type 'NR' if not reported): | text |
| | 154 | st_ci_level | If the confidence interval (CI) was not a 95% CI, specify the CI level (e.g. 90% CI, 99% CI); otherwise leave blank | text |
| | 155 | st_pvalue<br>Show the field ONLY if:<br>[st_data_avail] = '1' | Specify the exact P-value for the study effect estimate (type 'NR' if not reported): | text |
| | 156 | st_direction<br>Show the field ONLY if:<br>[st_data_avail] = '1' | Specify the direction of the study effect estimate (note: ignore the statistical significance of the effect, just focus on the direction): | radio<br>1 Favours intervention or exposure<br>2 Favours comparator<br>3 Neutral (e.g. RR is somewhere between 0.95 and 1.05 or MD is somewhere between -0.05 and 0.05) |

| | | | | |
|---|---|---|---|---|
| | | | | 4 Unclear |
| | 157 | st_location<br>Show the field ONLY if:<br>[st_data_avail] = '1' | Specify the location of the data in the report (e.g. Table 2, Figure 1, text page 378): | text |
| | 158 | incl_ma<br>Show the field ONLY if:<br>[st_data_avail] = '1' | Does the study effect estimate and 95% CI match the one included in the index meta-analysis? (note: if the study effect estimate and 95% CI were not reported in the study paper, calculate these values from the summary statistics) | radio<br>1 Yes<br>2 No<br>3 Unclear |
| | 159 | st_error<br>Show the field ONLY if:<br>[incl_ma] = '1' | Were the data entered by the systematic reviewers incorrect in any way (e.g. wrong sample size for cross-over trials, or standard error entered as standard deviation)? | yesno<br>1 Yes<br>0 No |
| | 160 | st_error_text<br>Show the field ONLY if:<br>[st_error] = '1' | Describe how the data were entered incorrectly by the systematic reviewers: | notes |
| | 161 | st_notes | Notes | notes |

| | | | | |
|---|---|---|---|---|
| | 162 | study_data_complete | Section Header: *Form Status*<br><br>Complete? | dropdown<br>0 Incomplete<br>1 Unverified<br>2 Complete |

**Supplementary Table S3: Content of decision rules reported in systematic review protocols and systematic reviews**

| Type of decision rule | Protocols (N=14) n (%) | Systematic reviews (N=42) n (%) |
|---|---|---|
| Measurement of outcome | | |
| Not applicable as the index outcome was defined by a particular /definition/criteria | 1 (7) | 1 (2) |
| No decision rule | 13 (93) | 41 (98) |
| Intervention/exposure and /control groups | | |
| If the same intervention/exposure, but more than one intake level, then select the highest/lowest level | 1 (7) | 19 (45) |
| If the same intervention/exposure, but with two intake levels, then combine into one group; | 1 (7) | 2 (5) |
| If two different interventions/ exposures, include each as separate comparison | 1 (7) | 2 (5) |
| *Other | - | 1 (2) |
| No decision rule | 11 (79) | 18 (43) |
| Time points | | |
| End of the study | 0 | 1 (2) |
| Other specific time point | 0 | 2 (5) |
| No decision rule | 14 (100) | 39 (93) |
| Final versus change from baseline values | | |
| Final values preferred | 0 | 0 |
| Change from baseline values preferred | 0 | 6 (14) |
| No decision rule | 14 (100) | 36 (86) |
| Unadjusted versus covariate adjusted analyses | | |
| Unadjusted analyses preferred to adjusted | 0 | 1(2) |
| Adjusted analyses preferred to unadjusted | 0 | 3 (7) |
| Most adjusted analyses preferred if multiple adjusted analyses were reported | 1 (7) | 3 (7) |
| No decision rule | 13 (93) | 35 (83) |
| Period or paired analyses in cross-over RCTs | | |
| First period results only | 0 | 2 (5) |
| No decision rule | 14 (100) | 40 (95) |
| Analyses undertaken on multiple samples | | |
| Intention-to-treat (ITT) analysis preferred | 0 | 0 |
| Modified ITT analysis preferred (if missing data, modified (last observation carried forward) ITT analysis preferred over per-protocol or as-treated analysis | 0 | 0 |
| Per protocol | 0 | 0 |
| As-treated analysis | 0 | 0 |
| No decision rule | 14 (100) | 42 (100) |

*Classified based on the authors' description (e.g. when two or more healthy dietary patterns were presented, researchers generally considered a healthy pattern one that mostly included protective foods, such as vegetables, fruits, fish, legumes, whole grains, and low-fat dairy products); ITT: Intention to treat; RCTs: Randomised Controlled Trials

**Supplementary Table S4: Number of systematic reviews that had a discrepancy between the review protocol and review in the eligibility criteria or decision rules to select results (N=14)**

| **Type of eligibility criterion and decision rule discrepancy** | **Any n (%)** | **Addition n (%)** | **Omission n (%)** | **Modification n (%)** |
|---|---|---|---|---|
| Total | | | | |
| Discrepancy in at least one eligibility criterion | 14 (100) | 14 (100) | 0 | 0 |
| Discrepancy in at least one decision rule | 14 (100) | 13 (93) | 0 | 1 (7) |
| Intervention/control groups | | | | |
| Eligibility criteria | 1 (7) | 1 (7) | 0 | 0 |
| Decision rule | 12 (86) | 11 (79) | 0 | 1 (7) |
| Time points | | | | |
| Eligibility criteria | 3 (21) | 3 (21) | 0 | 0 |
| Decision rule | 2 (14) | 2 (14) | 0 | 0 |
| Analyses | | | | |
| Eligibility criteria for any type of analysis | 14 (100) | 14 (100) | 0 | 0 |
| Decision rule for any type of analysis | 14 (100) | 14 (100) | 0 | 0 |
| Decision rule for analyses undertaken on multiple samples (e.g. ITT versus per-protocol) | 0 | 0 | 0 | 0 |
| Decision rule for unadjusted versus covariate adjusted analyses | 13 (93) | 13 (93) | 0 | 0 |
| Decision rule for period versus paired analyses in cross- over RCTs | 0 | 0 | 0 | 0 |
| Other decision rule | 0 | 0 | 0 | 0 |

ITT: Intention to treat; RCTs: Randomised Controlled Trials
